# Chest CT features of COVID-19 in the region of Abu Dhabi, UAE- A single institute study

**DOI:** 10.1101/2020.11.14.20229096

**Authors:** Ghufran Aref Saeed, Abeer Ahmed Al Helali, Safaa Almazrouei, Asad Shah, Luai A. Ahmed

## Abstract

**OBJECTIVE:** Our aim is to investigate high resolution CT features of COVID-19 infection in Abu Dhabi, UAE, and to compare the diagnostic performance of CT scan with RT-PCR test.

**METHODS:** Data of consecutive patients who were suspected to have COVID-19 infection and presented to our hospital, was collected from March 2, 2020, until April 12, 2020. All patients underwent RT-PCR test; out of which 53.8% had chest CT scan done. Using RT-PCR as a standard reference, the sensitivity and specify of CT scan was calculated. We also analyzed the most common imaging findings in patients with positive RT-PCR results.

**RESULTS:** The typical HRCT findings were seen in 50 scans (65.8%) out of total positive ones; 44 (77.2%) with positive RT-PCR results and 6 (31.6%) with negative results. The peripheral disease distribution was seen in 86%, multilobe involvement in 70%, bilateral in 82%, and posterior in 82% of the 50 scans.

The ground glass opacities were seen in 50/74 (89.3%) of positive RT-PCR group. The recognized GGO patterns in these scans were: rounded 50%, linear 38%, and crazy-paving 24%.

Using RT-PCR as a standard of reference, chest HRCT scan revealed sensitivity of 68.8% and specificity of 70%.

**CONCLUSION:** The commonest HRCT findings in patients with COVID-19 pneumonia were peripheral, posterior, bilateral, multilobe rounded ground glass opacities.

## INTRODUCTION

In December 2019, multiple patients were diagnosed with pneumonia of unknown cause in Wuhan, China. Afterwards, it was determined that a novel strain of coronavirus was responsible for the outbreak, which was related to a seafood wholesale market (1). The disease has quickly spread from Wuhan to other areas (2). On January 29, the Ministry of Health and Prevention (MoHAP) confirmed the UAE’s first case of COVID-19 disease (3). In early March 2020, the WHO declared this outbreak a global pandemic.

The disease presentation is variable, ranging from no symptoms to severe respiratory distress and even death. Abnormal radiological findings have been increasingly reported in the literature. CT scans of the chest have shown a high sensitivity and features which can be considered specific for COVID-19 pneumonia (4,5). On the other hand, many studies have shown variable sensitivity and specificity of chest scans with increasing false negative rates (6).

## METHODS

### Data collection

This retrospective cross-sectional study was approved by the Ethics Review Committee of Department of Health-Abu Dhabi, UAE. The informed consent was waved off as per the committee. We collected clinical and laboratory data for analysis, derived from an electronic medical record system, from March 2, 2020, until April 12, 2020, of patients who were suspected to have COVID-19 infection. Scan images were collected and evaluated using the Picture Archiving and Communication Systems (PACS).

### HRCT inspection

All chest HRCT scans were performed on the day of patients’ presentation using a VCT GE 64 scanner. Patients were placed in a supine position. Scanning parameters were: scan direction (craniocaudally), tube voltage (120KV), tube current (100-600 mA)-smart mA dose modulation, slice collimation (64 × 0.625 mm), width (0.625 × 0.625 mm), pitch (1), rotation time (0.5 s), scan length (60.00 – I300.00 s).

### HRCT image analysis

Two radiologists with more than 8 years of experience evaluated the images to identify the disease characteristics of viral pneumonia in each patient. The scans were first assessed whether negative or positive. Positive scans were further classified into typical, indeterminate and atypical classes according to the Radiological Society of North America (RSNA) Expert Consensus (7). The evaluated radiological features were: ground glass opacities GGO, different GGO patterns, presence of peripheral, bilateral, posterior, multilobe (>2) distribution, consolidation, lymphadenopathy, bronchiectasis, nodules surrounded by GGO, interlobular septal thickening, pericardial effusion, pleural effusion, and cavitation.

### Statistical analysis

Descriptive statistics of patients’ demographic, clinical, laboratory and imaging characteristics are reported as means (standard deviation (SD)) and numbers and relative frequencies. Continuous variables were compared using t-test and categorical variables will be compared using Chi-Square test or Fisher’s exact test. Using RT-PCR test results as gold standard, the sensitivity and specificity were calculated to estimate the diagnostic performance of chest HRCT images. The analysis was performed using STATA version 16.1 (Stata Corp, College Station, TX, USA), and p-value less than 0.05 defined statistical significance.

## Results

### Baseline information

Our population included 173 consecutive patients who were suspected to have COVID-19 infection. The infection was confirmed in 104 (60.1%) and excluded in 69 (39.9%) of the patients using RT-PCR as a gold standard test. Two nasopharyngeal RT-PCR tests were performed within 48 hours. Three patients who tested initially negative and had typical CT scan findings, tested positive on the repeated RT-PCR. Those patients were considered as confirmed COVID-19 cases.

The mean age was 38.6 ± 1.5 years in the RT-PCR positive group [73 men (70.2%), 31 women (29.8%)] and 39.5± 2.4 years in the negative one [41 men (59.4%), 28 women (40.6%)]. Risk factors were seen in 32 patients (30.8%) with positive RT-PCR. Recent travel history (within 30 days) and direct exposure to known COVID-19 patient were strongly associate with RT-PCR positive results (n=63, 60.6%, p-value <0.0001). 149/173 patients were symptomatic; in which 84 patients found to have positive RT-PCR results. Common presenting symptoms in this group were fever, dry cough and shortness of breath [n=56 (53.9%, p-value *0.001*), n=48 (46.2%, p-value *0.001*) and n=27 (26.0%), respectively] (Table 1).

**Table 1:** Characteristics of baseline data for suspected and confirmed COVID-19 infected patients.

|  | **All patients** | **PCR positive patients** | **PCR negative patients** | **p-value** |
| --- | --- | --- | --- | --- |
| N | 173 (100.0) | 104 (60.1) | 69 (39.9) |  |
| **Risk factors** |  |  |  |  |
| Hypertension | 22 (12.7) | 11 (10.6) | 11 (15.9) | 0.300 |
| Diabetes mellitus | 22 (12.7) | 11 (10.6) | 11 (15.9) | 0.300 |
| CAD | 9 (5.2) | 4 (3.9) | 5 (7.3) | 0.486 |
| Renal disease | 5 (2.9) | 0 (0.0) | 5 (7.3) | 0.009 |
| Immunocompromised | 8 (4.6) | 3 (2.9) | 5 (7.3) | 0.181 |
| Smoking | 11 (6.4) | 9 (8.7) | 2 (2.9) | 0.203 |
| Asthma | 7 (4.1) | 6 (5.8) | 1 (1.5) | 0.245 |
| Other lung diseases | 3 (1.7) | 1 (1.0) | 2 (2.9) | 0.564 |
| Other health problems | 31 (17.9) | 19 (18.3) | 12 (17.4) | 0.883 |
| History of travel/Exposure | 78 (45.1) | 63 (60.6) | 15 (21.7) | <0.0001 |
| **Symptoms** |  |  |  |  |
| Any Symptom | 149 (86.1) | 84 (80.8) | 65 (94.2) | 0.012 |
| Fever | 75 (43.4) | 56 (53.9) | 19 (27.5) | 0.001 |
| SOB | 40 (23.1) | 27 (26.0) | 13 (18.8) | 0.277 |
| Chest pain | 8 (4.6) | 7 (6.7) | 1 (1.5) | 0.147 |
| Runny nose | 16 (9.3) | 11 (10.6) | 5 (7.3) | 0.595 |
| Dry cough | 63 (36.4) | 48 (46.2) | 15 (21.7) | 0.001 |
| Productive cough | 19 (11.0) | 10 (9.6) | 9 (13.0) | 0.480 |
| Sore throat | 31 (17.9) | 24 (23.1) | 7 (10.1) | 0.030 |
| Diarrhea | 13 (7.5) | 12 (11.5) | 1 (1.5) | 0.014 |
| Nausea or vomiting | 12 (6.9) | 11 (10.6) | 1 (1.5) | 0.021 |
| Body aches/fatigue | 24 (13.9) | 18 (17.3) | 6 (8.7) | 0.109 |
| Other symptoms | 42 (24.3) | 37 (35.6) | 5 (7.3) | <0.0001 |

- Numbers listed (in brackets) represent percentages. - All percentages are column percent.

Laboratory results for patients with RT-PCR positive results showed lymphopenia (normal value 1.5-4⍰×⍰109/L) in 43 patients (41.4%), elevated CRP (normal value ≤5 mg/L) in 55 patients (51.9%), high d-dimer (normal value ≤0.5 mcg/mL) in 35 patients (33.7%), elevated serum amylase (normal value 28-100 units/L) and lipase (normal value 13-60 IU/L) n= 11 patients (10.6%).

### HRCT evaluation

Excluding one scan due to significant motion artefact; total of 93 patients (53.8%) who had HRCT scan done were included in the assessment. The scans were positive in 74 patients (79.6%) and negative in 19 patients (20.4%) (Figure1).

**Figure1:**
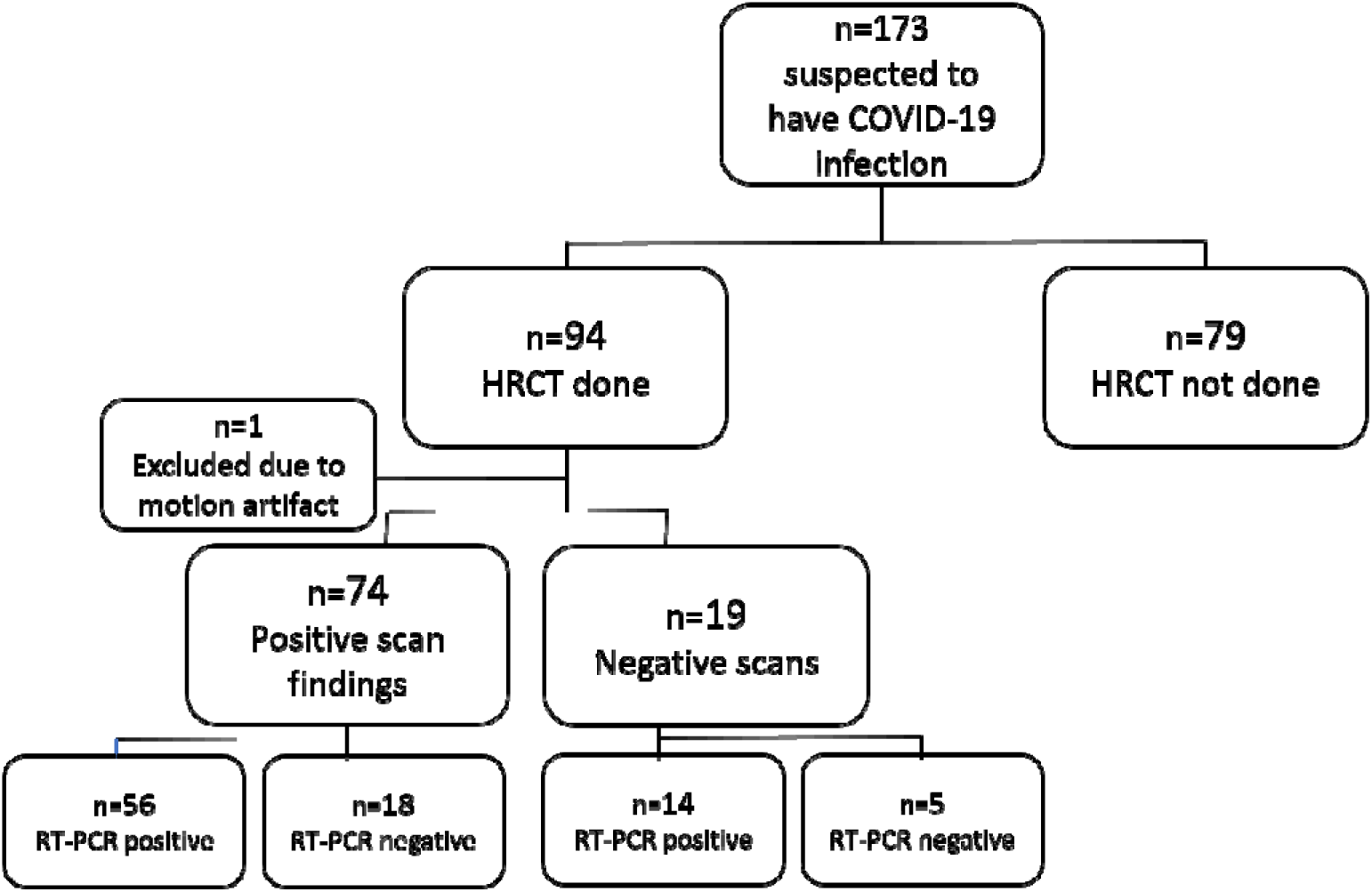
Cohort selection and distribution. The positive CT findings were classified into typical, indeterminate and atypical according to the Radiological Society of North America (RSNA) Expert Consensus. Typical findings were seen in 68% of positive scans, indeterminate in 12% and atypical in 20% (Figure2). The findings in the RT-PCR positive group were: typical n= 44 (77.2%), indeterminate n= 6 (10.5%), and atypical n=6 (10.5%). In RT-PCR negative group: typical n= 6 (31.6%), indeterminate n=3 (15.8%), and atypical n=9 (47.4%). It is worth to mention that 14/19 of the patients with negative scan results, tested positive by RT-PCR.

**Figure2:**
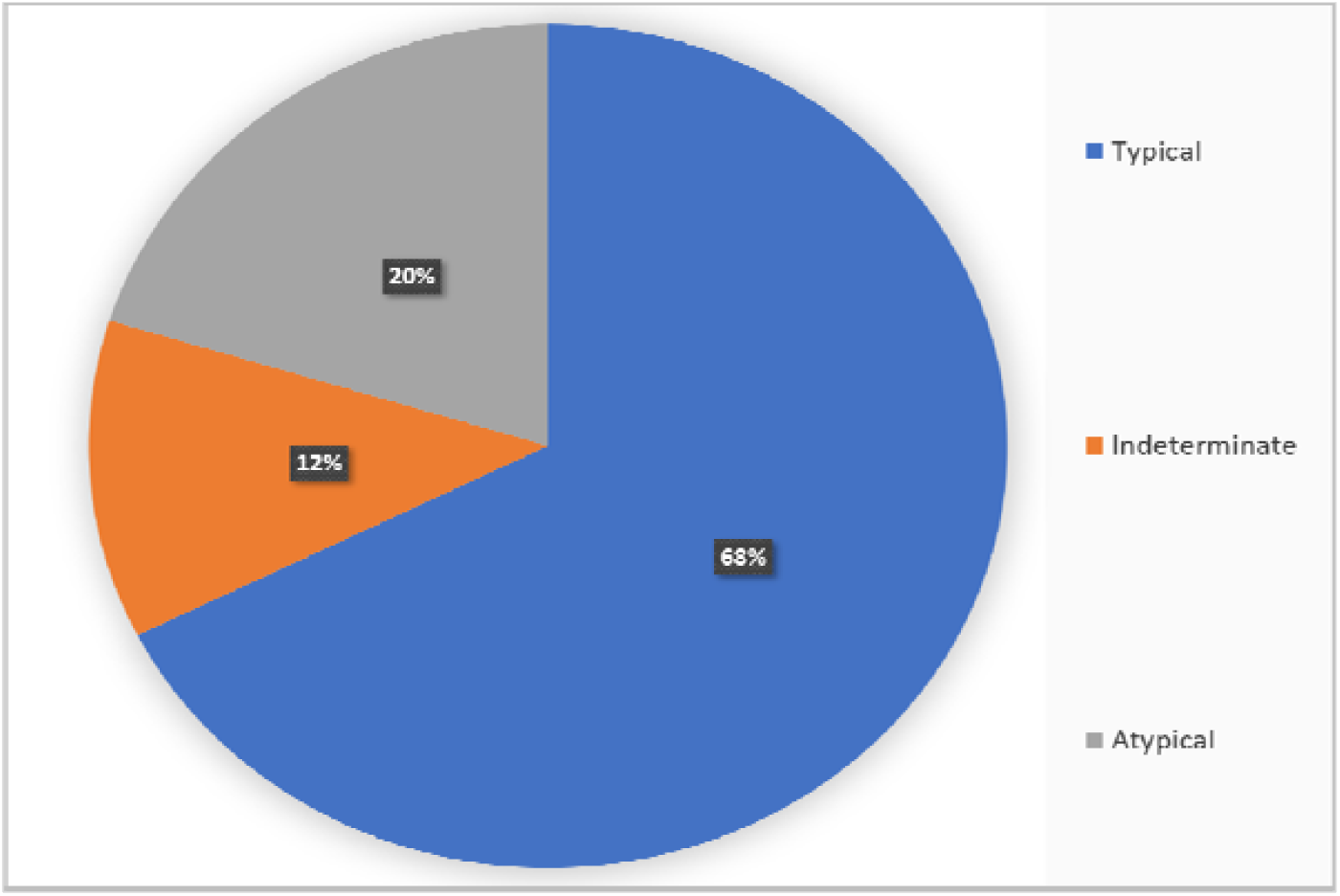
Classified general HRCT results. Ground glass pattern was seen in 59 scans out of 74 positive ones. 50/74 scans (89.3%) were for patients with positive RT-PCR results, and 9/74 were for patients with negative RT-PCR results. Out of the 50 scans showing GGO, the recognized patterns were: rounded 50% (36.2-63.8), linear 38% (25.4-52.4) and crazy-paving 24% (14.0-38.1), (Figure 3). Although the linear patter was the second most common pattern seen in diseased patients, it can be non-specific as was also seen in 7 patients (38.9%) who tested negative by RT-PCR. The peripheral disease distribution was seen in 86% (73.0-93.3), multilobe involvement in 70% (55.7-81.3), bilateral in 82% (68.5-90.5), posterior in 82% (68.5-90.5), nodules surrounded by GGO in 0%, interlobular septal thickening in 42% (28.9-56.3), consolidation in 12% (5.4-24.6), bronchiectasis in 2% (0.3-13.5), pericardial and pleural effusion in 0%, and cavitation in 2% (0.3-13.5), (Table 2).

**Figure 3:**
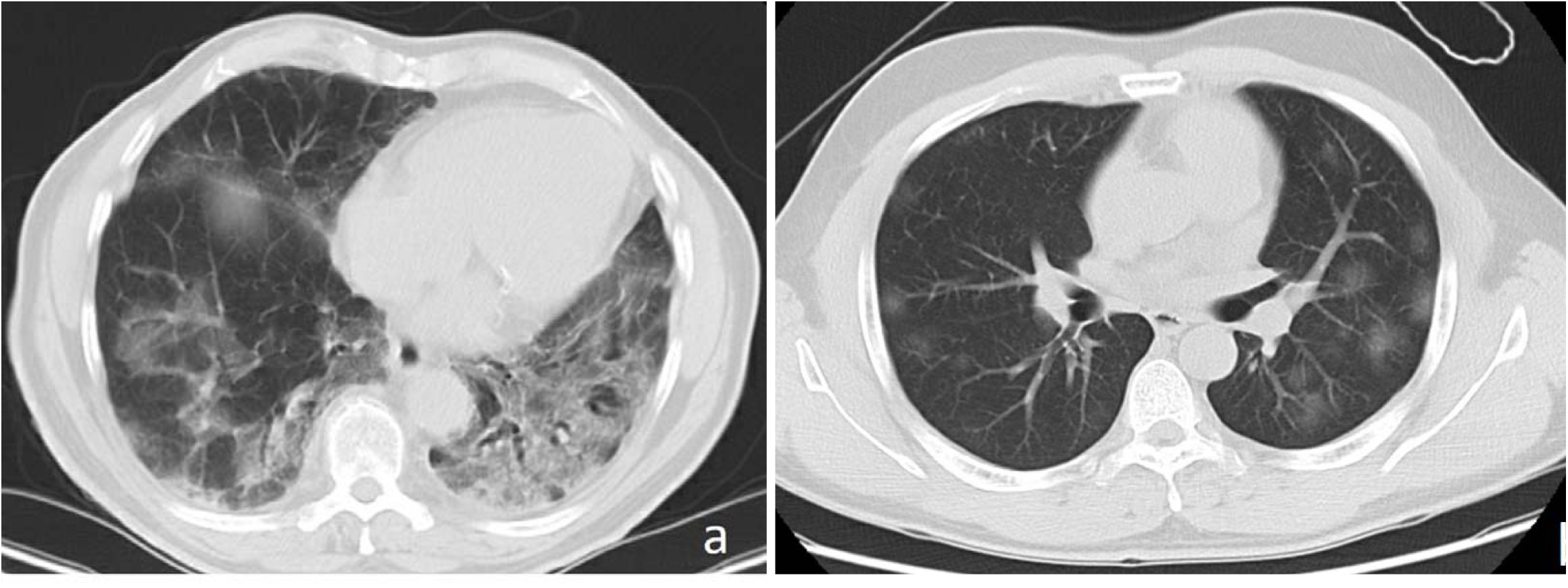
Axial thin-sections of unenhanced CT scan of two patients with COVID-19 pneumonia (a) Scan shows bilateral ground glass opacities with septal thickening (crazy paving pattern) (b) Scan shows bilateral ground glass opacities with rounded morphology.

**Table 2:**
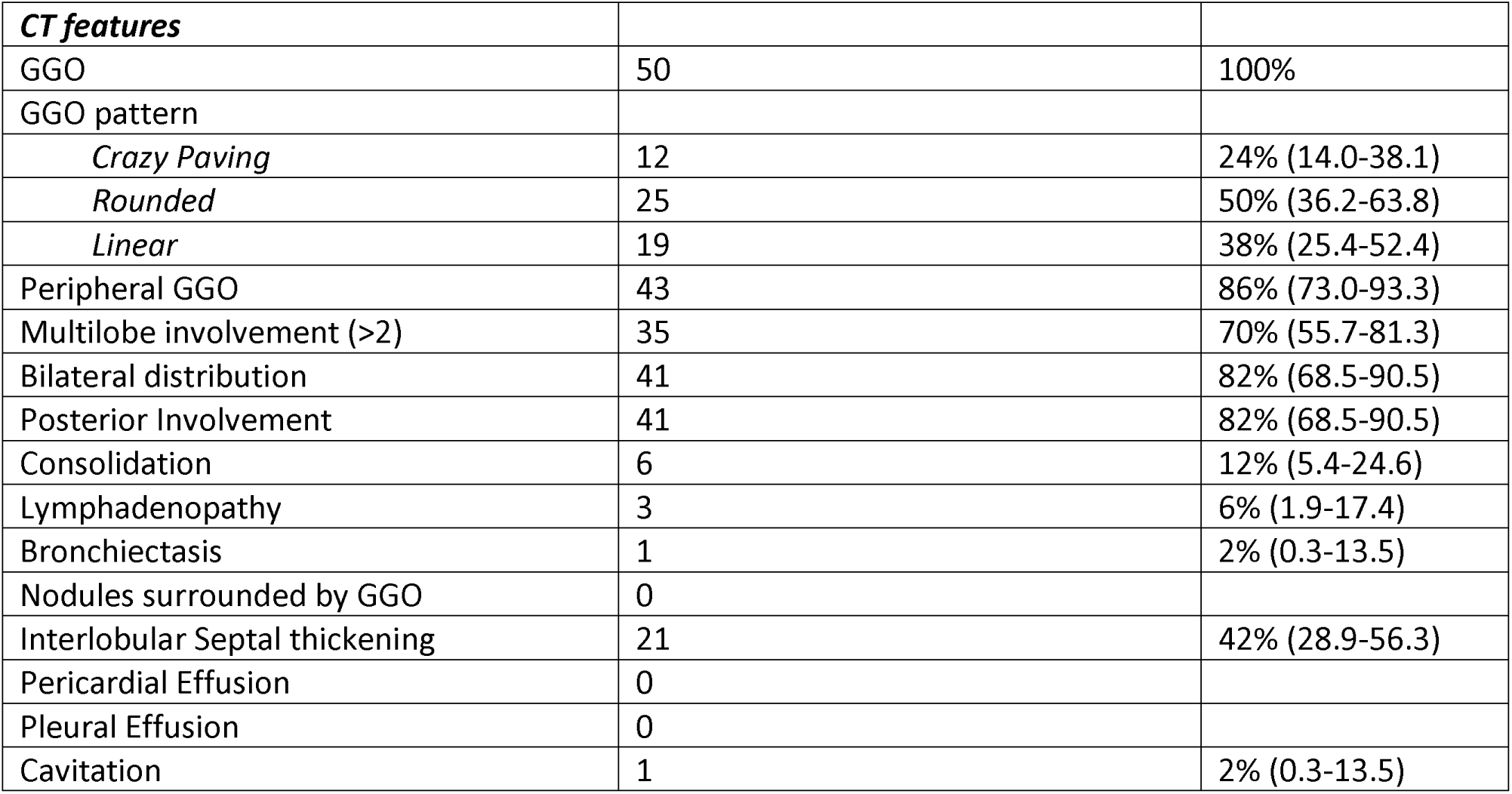
CT features in 50 patients with PCR confirmed COVID-19.

#### CT Diagnostic Performance

The diagnostic performance of CT including the sensitivity and specificity was calculated based on typical and atypical CT features for COVID-19 infection, using RT-PCR as a standard of reference. The results showed a **sensitivity** of 68.8% (95% CI 55.94% to 79.76%), **specificity** of 70% (95% CI 45.72% to 88.11%), and **accuracy** of 69.05% (CI 58.02% to 78.69%), (Table 3).

**Table 3:**
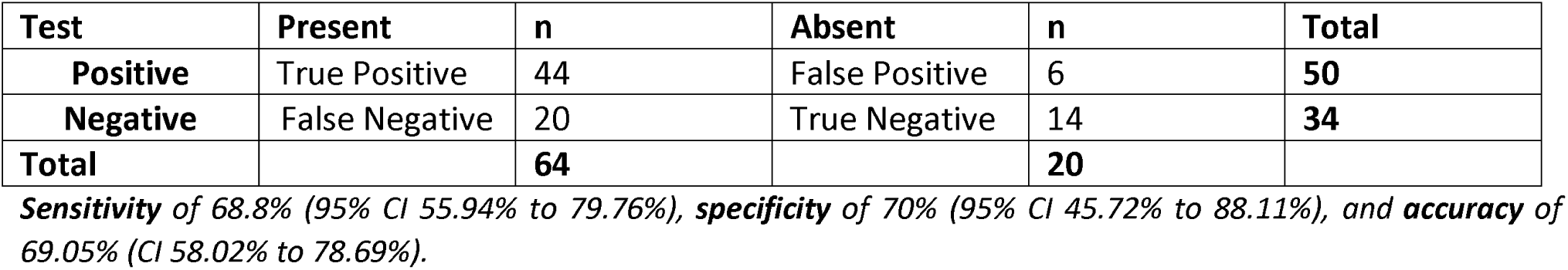
Diagnostic performance of HRCT using RT-PCR as gold standard.

| Test | Present | n | Absent | n | Total |
| --- | --- | --- | --- | --- | --- |
| **Positive** | True Positive | 44 | False Positive | 6 | **50** |
| **Negative** | False Negative | 20 | True Negative | 14 | **34** |
| **Total** |  | **64** |  | **20** |  |

**Sensitivity** of 68.8% (95% CI 55.94% to 79.76%), **specificity** of 70% (95% CI 45.72% to 88.11%), and **accuracy** of 69.05% (CI 58.02% to 78.69%).

## Discussion

Symptoms in patients with COVID-19 infection are usually developed in response to the direct viral destruction of lung epithelial cells or T-cell mediated immunological response (8). In this study, and in accordance with the systematic review performed by Grant *et al*. of 24,410 adults with confirmed COVID-19 infection from 9 countries; fever (53.9% vs 78%), and cough (55.8% vs 57%) were the most prevalent symptoms (9).

Moreover, laboratory results can reflect the general effect of the disease in the body. CRP can be elevated in multiple conditions, like infection and inflammation (10). It has been suggested that raised CRP and d-dimer levels are linked to a poor outcome in patients with COVID-19 disease. (11) Similarly, lymphopenia can be a good indicator of disease severity (12). Virus-related pancreatic injury with rising pancreatic enzymes and even pancreatitis were also described in the literature (13,14).

Regarding the CT findings, our results were comparable to the systemic review of 919 patients done by Salehi *et al*. (15). The ground glass opacities were found in (89.3% vs 88%), pulmonary consolidations (12% vs 31%), peripheral GGO distribution (86% vs 76%), bilateral involvement (82% vs 87.5%), and multilobe involvement (70% vs 78.8%).

The ground glass opacity appears as a mild increase in lung density due to pulmonary interstitial thickening or partial filling of the alveoli (16,17). Multiple studies have further characterized the GGO pattern in patients with COVID-19 pneumonia (18). The crazy-paving pattern and consolidation were more common in later stages of the disease (19). Although GGO can be seen in various pathologies, its pattern and distribution along with the clinical picture can favor one diagnosis over the other. The typical findings in patients with negative RT-PCR results can be attribute to other pathologies mimicking the typical CT appearance; such as influenza pneumonia, organizing pneumonia like in drug toxicity and connective tissue disease (20). Recently, new publications aim to differentiate Influenza A from COVID-19 pneumonia by identifying specific CT imaging features (21,22). In our analysis, among the six patients who were found to have typical CT findings for COVID-19 pneumonia but negative RT-PCT results; one patient was confirmed to have Influenza A pneumonia (Figure4). Another had Mycoplasma pneumonia with background of cardiogenic pulmonary edema (Figure5).

**Figure 4:**
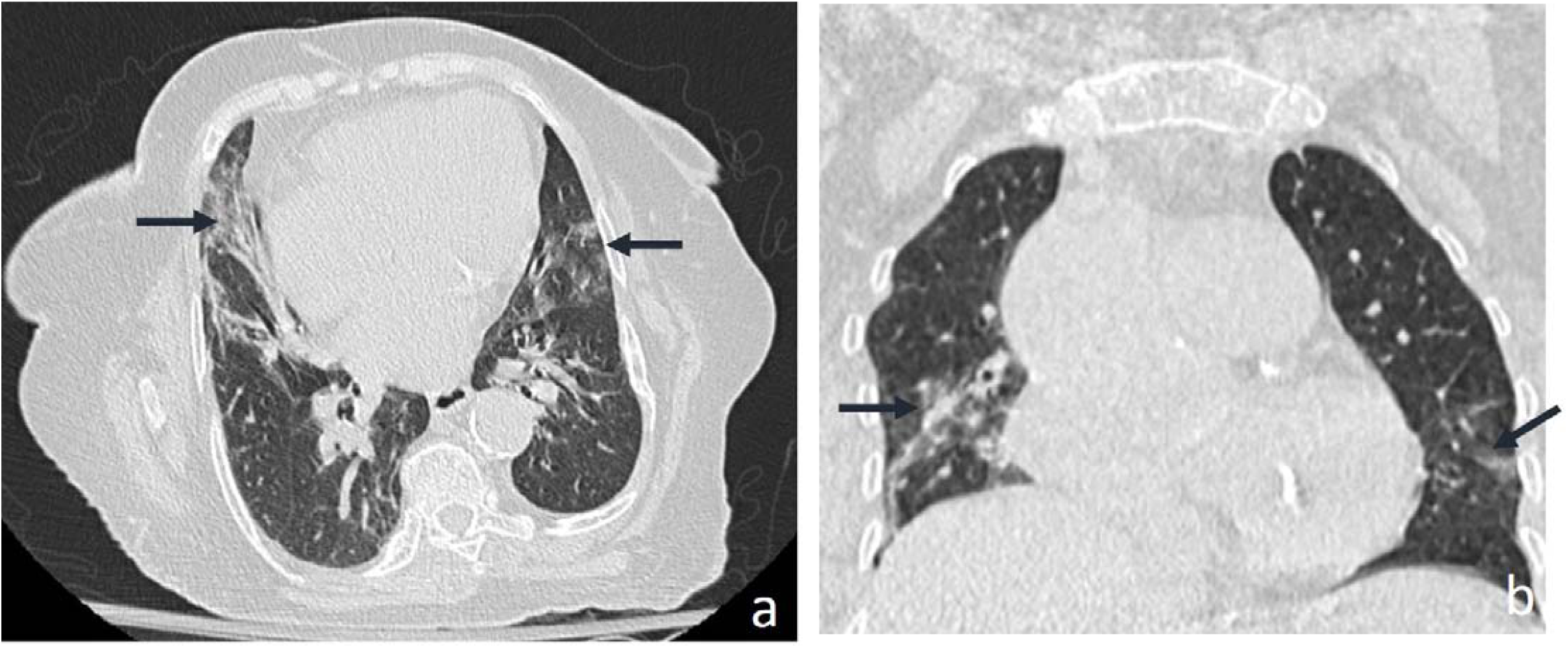
Axial (a) and coronal (b) thin-sections of unenhanced CT scan which was reported as typical for COVID-19 pneumonia. Note the bilateral peripheral lower lobe linear GGO (arrows). The patient was confirmed to have Influenza A pneumonia and his two RT-PCR tests were negative for COVID-19 pneumonia.

**Figure 5:**
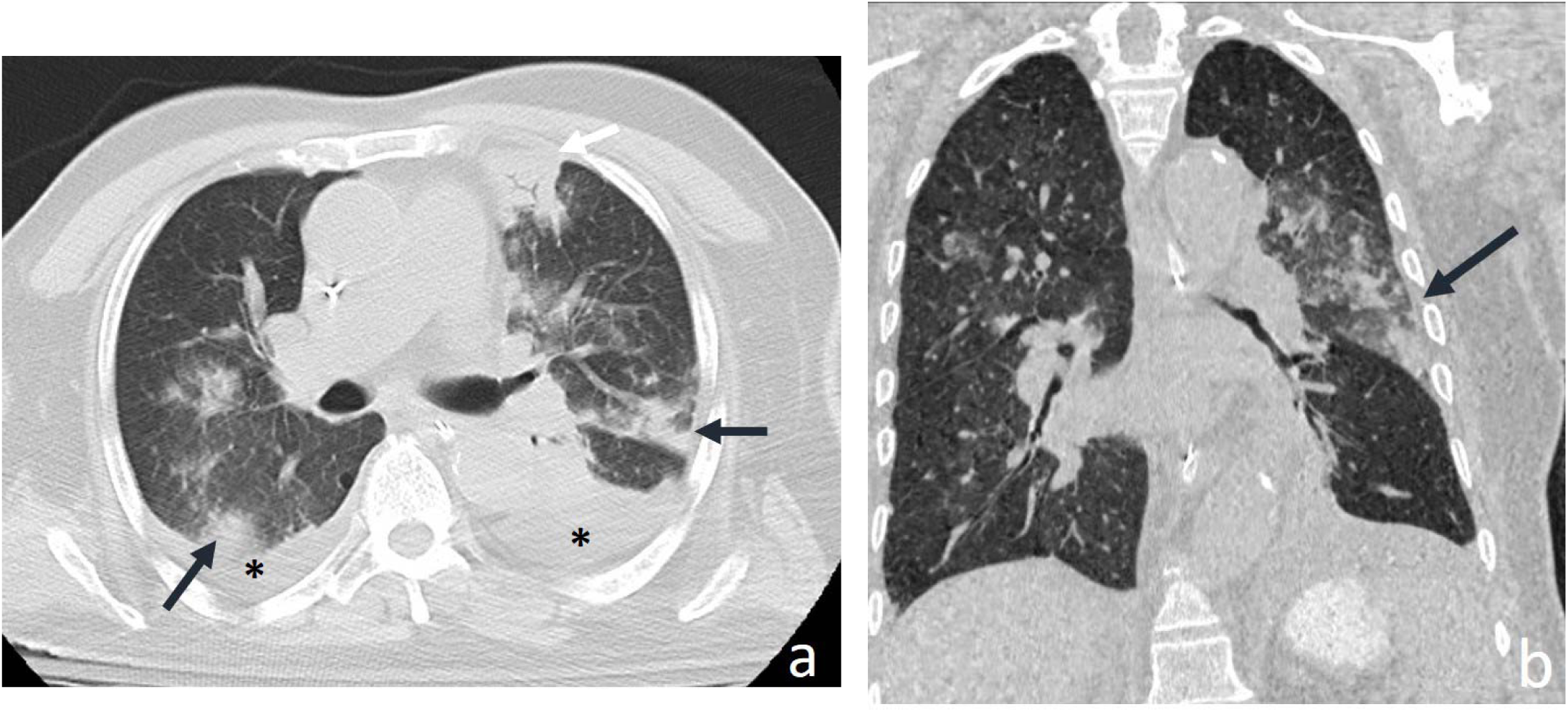
Axial (a) and coronal (b) thin-sections of unenhanced CT scan which was reported as combination of typical and atypical patterns for COVID-19 pneumonia (concurrent pathologies). Note the bilateral peripheral GGO (black arrows), consolidation (white arrow), and bilateral pleural effusion (asterix). The patient was known to have heart failure and confirmed to have Mycoplasma pneumonia. His two consecutive RT-PCR tests were negative for COVID-19 pneumonia.

Remarkably, a third patient who presented with dry cough, SOB, lymphopenia, elevated CRP and d-dimer, tested negative for COVID-19 infection by RT-PCR initially at the time of CT scan. The repeated test after 3 days showed a positive result. Furthermore, one patient was asymptomatic and the other two patients, although were mildly symptomatic, have no follow up information in our records. Cases with distinguishing imaging features for COVID-19 pneumonia seen on CT scans in asymptomatic patients have been reported (23)

On the other hand, and in spite of the relatively high false negative rate (n=14, 20%) which is seen in our analysis, this can still be explained by several factors. One is the young patient cohort, which is probably related to the high prevalence of immigrant workers in our region presenting with mild symptoms. Another aspect is the early large prompt screening program that was initiated in the United Arab Emirates. This brings the discussion forward on how these factors can affect the diagnostic performance of CT scan when compared with RT-PCR test.

Multiple studies have reported the sensitivity and specificity of CT scan in diagnosing COVID-19 pneumonia. The results were variable. Isikbay *et al*. have analyzed and described chest CT findings in patients with COVID-19 infection aboard the “Diamond Princess” cruise ship. A low sensitivity of 61% and 20% false negative rate in symptomatic patients were reported (24,25,26). This supports the European and American societies’ consensus, recommending that CT scan should not be used to screen for or as a first-line test to diagnose COVID-19 disease (27). The authors also suggested that sensitivities differ based on the selected cohort and the patient’s disease stage at which imaging was done. This heterogeneity can be also related to the experience of the radiologists and the severity of the epidemic, explaining the higher sensitivity values in Wuhan (28). Moreover, CT sensitivity and specificity can be highly affected by the adopted CT positivity threshold (29). A lower threshold would increase the sensitivity in expense of specificity, and vice versa.

Our study has some limitations. The small cohort and the fact that the sample was taken during early stages of the disease, just before the peak of the epidemic in our region, might have influenced the results. Moreover, the results may be biased by the use of the RT-PCR test as a standard of reference. Some studies have suggested that RT-PCR test carries false negative and positive rates (30,31). Additionally, the diagnostic performance of CT scan varies depending on the chosen threshold. Finally, histopathologic results from lung biopsies were not available to be correlated with imaging findings.

In summary, the study showed variable imaging patterns of COVID-19 disease affecting the lungs. The peripheral, posterior, bilateral, multilobe rounded ground glass opacities, were the commonest features seen in patients with COVID-19 pneumonia in the region of Abu Dhabi, UAE. The diagnostic performance of chest CT scan can be variable based on multiple factors.

## Data Availability

All data available among request by  the Institutional Review Board (IRB) and Department of health (DOH), Abu Dhabi, United Arab Emirates (UAE)

## Declaration of Competing Interest

No conflict of interest needs to be disclosed

## Funding sources

This research did not receive any specific grant from funding agencies in the public, commercial, or not-for-profit sectors.

## Abbreviations

COVID-19: (coronavirus disease)
CRP: (C-reactive protein)
CT: (computed tomography)
GGO: (ground-glass opacity)
HRCT: (high-resolution computed tomography)
PACS: (picture archiving and communication systems)
RT-PCR: (reverse transcription polymerase chain reaction)
CAD: (coronary artery disease)
SOB: (shortness of breath)
WHO: (World Health Organization)

## Notes

### Competing Interest Statement

The authors have declared no competing interest.

### Author Declarations

Institutional Review Board (IRB) and Department of health (DOH), Abu Dhabi, United Arab Emirates (UAE)

